# Negative impact of hyperglycemia on Tocilizumab therapy in COVID-19 patients

**DOI:** 10.1101/2020.04.29.20076570

**Authors:** Raffaele Marfella, Pasquale Paolisso, Celestino Sardu, Luca Bergamaschi, Emanuela Concetta D’Angelo, Michelangela Barbieri, Maria Rosaria Rizzo, Vincenzo Messina, Paolo Maggi, Nicola Coppola, Carmine Pizzi, Mauro Biffi, Pierluigi Viale, Nazzareno Galié, Giuseppe Paolisso

## Abstract

Tocilizumab is used for treating moderate-severe Covid-19 pneumonia by targeting IL-6 receptors (IL-6R) and reducing cytokine release, but the pooled rate ratio among diabetic patients with adverse vs those with the more favorable course was 2.26. To date, the hyperglycemia has been shown to increase IL-6 and IL-6R, which has been suggested as a severity predictor in lung diseases of Covid-19 patients. However, there are no data about the effects of tocilizumab therapy on outcomes of hyperglycemic Covid-19 patients with pneumonia. To investigate this unsolved need, 475 Covid-19 positive patients were retrospectively studied since March 1st, 2020. Among them, 78 patients with pneumonia disease and treated with tocilizumab were further evaluated for a severe outcome (encompassing both the use of mechanical ventilation and/or death). Thirty-one (39.7%) hyperglycemic and 47 (60.3%) normoglycemic Covid-19 positive patients (blood glucose levels >140 mg/dl, at admission and/or during hospital stay) were evaluated. Noteworthy, 20 (64%) of hyperglycemic and 11 (23.4%) of normoglycemic patients were also diabetics (P<0.01). At admission, more elevated IL-6 levels in hyperglycemic patients were found and persists even after Tocilizumab administration. In a risk adjusted Cox-regression analysis, Tocilizumab in hyperglycemic did not attenuate the risks of severe outcome as did in normoglycemic patients (p<0.009). Therefore, we could conclude that reduced effects of Tocilizumab in hyperglycemic patients may due to the higher plasma IL-6 levels. Interestingly, when we added IL-6 levels in a Cox regression model the significance for the tocilizumab effect was lost (p<0.07). In this context, our observations evidence that optimal Covid-19 infection management with tocilizumab is not achieved during hyperglycemia both in diabetic and non-diabetic patients.

---

To calm inflammatory storm, tocilizumab was used for treating moderate-severe Covid-19 pneumonia by targeting IL-6 receptors (IL-6R) and reducing cytokine release **(1)**. Despite the optimal management of Covid-19 infection including tocilizumab, the pooled rate ratio among diabetic patients with adverse vs those with the more favorable course was 2.26 (**2**). An important factor in any form of infection control in patients with diabetes seems to be the glucose control (**2**). Hyperglycemia has been shown to increaseIL-6 and IL-6R (**3**), which has been suggested as a severity predictor in lung diseases of Covid-19 patients **(1)**. However, there are no data about the effects of tocilizumab therapy on outcomes of hyperglycemic Covid-19 patients with pneumonia. To investigate this unsolved need, 475 Covid-19 positive patients admitted to Infection Disease Departments (University of Bologna, University Vanvitelli Napoli, and San Sebastiano Caserta Hospital, Italy) since March 1^st^, 2020 were retrospectively studied. The study was approved by the Ethical Committee of The Institutional Review Boards. The patients and patients’ families signed informed consent to participate in the study. The diagnosis of COVID-19 was established according to World Health Organization interim guidance and confirmed by RNA detection of the SARS-CoV-2 in the microbiology laboratory of the hospitals. Among them, only 78patients with pneumonia disease and treated with tocilizumab were further evaluated for a severe outcome (encompassing both the use of mechanical ventilation and/or death). Thirty-one (39.7%) hyperglycemic and 47 (60.3%) normoglycemic Covid-19 positive patients (blood glucose levels ≥140 mg/dl, at admission and/or during hospital stay) were evaluated (**4**). Noteworthy, 20 (64%) of hyperglycemic and 11 (23.4%) of normoglycemic patients were also diabetics (P<0.01). Admission glucose levels were 187±48 vs 103+23 mg/dl (p<0.01) in hyperglycemic vs in normoglycemic patients. Along with the hospitalization, mean glucose levels were 157±15 vs 122 ±12 mg/dl (p<0.01) in hyperglycemic vs normoglycemic patients. At admission, more elevated IL-6 levels in hyperglycemic patients were found and persists even after Tocilizumab administration (**Figure-A)**. In a risk adjusted Cox-regression analysis, Tocilizumab in hyperglycemic did not attenuate the risks of severe outcome as did in normoglycemic patients (**p<0.009**) (**Figure-B)**.

**Figure A.**
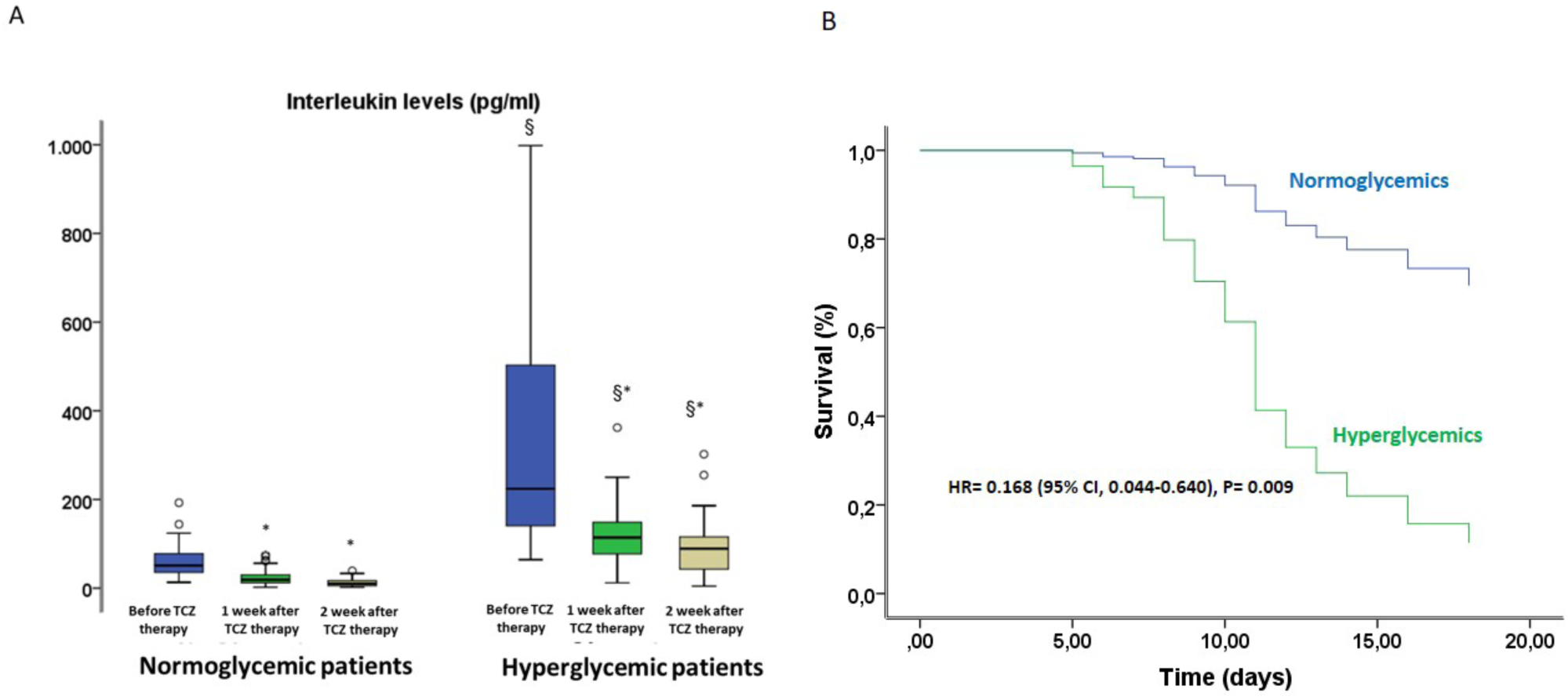
All patients were treated with intravenous (IV) infusion of tocilizumab, dosed at 8 mg/kg. Seven hyperglycemic patients (22.6%) and 5 normoglycemic patients (10.6%) received 2 IV infusion of tocilizumab (P = 0.134). The mean±SD of age was 65.7±13.4 years in hyperglycemic and 66.6±12.2 in normoglycemic patients (P= 0.662), 21 (61.8%) of hyperglycemic and 34 (72.3%) of normoglycemic patients were male (P=0.425). The median time from illness onset (before admission) to discharge or death was 17.6±7.2 days in hyperglycemic and 18.1±6.6 days in normoglycemic patients. Nineteen hyperglycemic (61.3%) and 26 normoglycemic (55.3%) patients were hypertensive (P= 0.388), 10 hyperglycemic (32.3%) and 7 normoglycemic (14.9%) patients were dyslipidemics (P= 0.063), 6 hyperglycemic (19.4%) and 15 normoglycemic (31.9%) patients were smokers (P= 0.168). There was no significant difference in blood pressure, creatinine and troponin levels between hyperglycemic and normoglycemic patients. All patients were treated with the standard protocol as antiviral treatment, tocilizumab and hydroxychloroquine. **Panel A)** IL-6 levels (ELISA KiT, RD System) before and after tocilizumab (TCZ) therapy in hyperglycemic and normoglycemic patients. Boxplot, a plot type that displays the median, 25th, and 75th percentiles and range. *P<0.05 vs before TCZ administration, §P<0.05 vs normoglycemic patients.

**Figure B.**
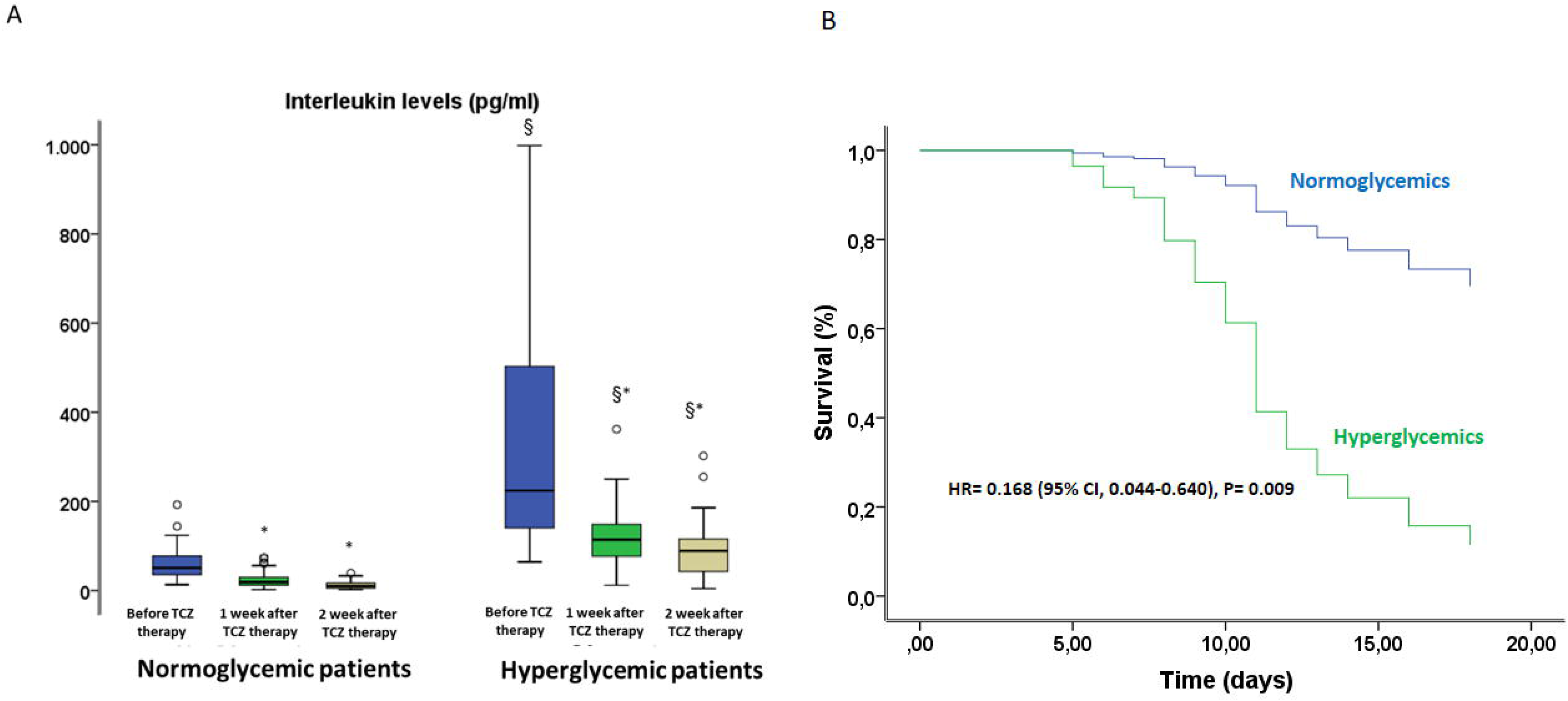
**Panel B)**Risk adjusted Cox-regression analysis curves showing survival from severe disease through 18 days for Covid-19 patients stratified by hyperglycemic vs normoglycemic patients. Cox models were adjusted for; BMI, age, gender, blood pressure heart rate, cholesterol, HDL-cholesterol, LDL-cholesterol, triglycerides levels, troponin levels, heart diseases, hypertension, dyslipidemia, current smoking, beta-blockers, ace-inhibitors, calcium inhibitors, thiazide diuretics, aspirin. All statistical analyses were performed with SPSS, version 23.0 (IBM Corp) for Windows. A 2-sided P <0.05 was considered statistically significant.

The reduced effects of Tocilizumab in hyperglycemic patients may due to the higher plasma IL-6 levels. Previous study evidenced that tocilizumab efficacy was inversely proportional to the baseline IL-6 levels in patients with rheumatoid arthritis (**5**). Accordingly, our data evidence that hyperglycemic patients had IL-6 levels at admission 5-fold higher as compared to the normoglycemic patients. Interestingly, when we added IL-6 levels in a Cox regression model the significance for the tocilizumab effect was lost (p<0.07). In this context, our observations evidence that optimal Covid-19 infection management with tocilizumab is not achieved during hyperglycemia both in diabetic and non-diabetic patients. Thus, these data may be of interest for ongoing clinical trials on tocilizumab effects on Covid-19 patients and for the optimal control of glycemia in this subset.

## Data Availability

Data are available.

